# A genome-wide association study of problematic sexual behaviour: genetic overlap with psychiatric, behavioural and personality phenotypes

**DOI:** 10.64898/2026.05.15.26351052

**Authors:** Jerome Clifford Foo, Shui Jiang, Yaroslav Ilnytskyy, Dongping Li, Xiuying Hu, Randolph Arnau, Rick Isenberg, Bradley Green, Igor Kovalchuk, Josef Frank, Rohit Lodhi, Leslie Roper, David Wishart, BASIC, Fabian Streit, Patrick J Carnes, Katherine Jean Aitchison

**Affiliations:** Department of Psychiatry, College of Health Sciences, University of Alberta, Edmonton, AB, Canada; Neuroscience and Mental Health Institute, University of Alberta, Edmonton, AB, Canada; Department of Genetic Epidemiology in Psychiatry, Central Institute of Mental Health, Medical Faculty Mannheim, University of Heidelberg, Mannheim, Germany; Biological Sciences, Faculty of Arts & Science, University of Lethbridge, Lethbridge, AB, Canada; School of Psychology, University of Southern Mississippi, Hattiesburg, MS, USA; Psychological Counselling Services, Ltd., Scottsdale, AZ, USA; Department of Psychology and Counseling, University of Texas at Tyler, Tyler, TX, USA; Department of Psychiatry, Schulich School of Medicine and Dentistry, Western University; Department of Biological Sciences, University of Alberta, Edmonton, AB, Canada; The Metabolomics Innovation Centre (TMIC), Edmonton, AB, Canada; Department of Computing Science, University of Alberta, Edmonton, AB, Canada; Department of Laboratory Medicine and Pathology, University of Alberta, Edmonton, AB, Canada; Faculty of Pharmacy and Pharmaceutical Sciences, University of Alberta, Edmonton, AB, Canada; Behavioral Addictions Studies and Insights Consortium*; Hector Institute for Artificial Intelligence in Psychiatry, Central Institute of Mental Health, Medical Faculty Mannheim, Heidelberg University, Mannheim, Germany; Department of Psychiatry and Psychotherapy, Central Institute of Mental Health, Medical Faculty Mannheim, Heidelberg University, Mannheim, Germany; German Center for Mental Health (DZPG), Partner Site Mannheim - Heidelberg - Ulm, Germany; The Meadows, Wickenburg, USA; Department of Medical Genetics, College of Health Sciences, University of Alberta, Edmonton, AB, Canada; Psychiatry Section, Division of Clinical Sciences, Northern Ontario School of Medicine, Thunder Bay, ON, Canada

## Abstract

Problematic Sexual Behaviour (PSB) is defined as difficult to control recurrent sexual behaviours that continue despite adverse consequences, leading to social and functional impairment. There is debate whether PSB is a disorder of compulsion or addiction; PSB often co-occurs with neuropsychiatric disorders, but further elucidation regarding underlying biology is required. A deficiency in reward neurotransmitter systems (reward deficiency syndrome: RDS) may underlie a shared vulnerability to addiction.

We conducted the first case-control genome wide association study (GWAS) of PSB in patients (n=448), and comparison participants with (n=196) and without PSB (n=1488). We used polygenic risk scores (PRS) to test genetic overlap with related psychiatric, behavioural and personality phenotypes. Three models were used: 1) All-PSB (patient + comparison) vs. controls, 2) Patient-PSB vs controls, and 3) RDS (yes/no).

Results suggested genetic overlap of PSB with psychiatric conditions, with PRS for major depression, substance use, and others associated with PSB status. PRS for related behavioural phenotypes (e.g., externalizing, age at first sex, number of lifetime sexual partners) and personality traits were also associated with PSB. The patient model showed stronger associations than the All-PSB model, and RDS had both shared and distinct genetics with PSB. As expected with the sample size, only suggestive hits were observed with single variant and gene-based tests.

PSB may share genetic mechanisms with various conditions. Further research in larger cohorts is needed to better understand the underlying genetics and environmental factors involved, and to improve diagnostic classification, intervention and treatment prospects.

## INTRODUCTION

Problematic sexual behavior (PSB) is broadly defined by the inability to control sexual urges and the continuation of such behaviors despite adverse consequences, which may lead to marked distress or social and functional impairment, as well as potential harm to oneself or others [1–3]. The estimated prevalence of PSB is as high as 7-10% and increasing [4], and may be higher in certain populations; due to a lack of consensus about its specific definition and diagnosis, estimates of prevalence in the population vary [5].

The heterogeneity in presentation and range of behaviours observed, overlap with other disorders, as well as unclear underlying mechanisms contribute to the uncertainty about PSB. On one hand, it is proposed that PSB is a type of impulse control disorder; compulsive sexual behaviour disorder (CSBD) was recently included in the International Statistical Classification of Diseases [6]. It has also been argued that PSB is a manifestation of a behavioural addiction [7]. PSB shows comorbidity with attention deficit hyperactivity disorder (ADHD) (22.6%) [8, 9] and this association may be driven by a complex combination of factors including impulsivity, emotional dysregulation, increased sexual desire and compulsivity, social and relationship difficulties, attachment issues, and increased stress leading to sexual behavior as a coping mechanism [10]. PSB is also observed to be comorbid with mood disorders such as depression, as well as with substance use disorders (SUD) [11]. These comorbidities may suggest that PSB is not confined to the boundaries of specific categories; clarifying the biological underpinnings represents a key step towards understanding the nature of PSB and informing the development of effective therapies and interventions.

A complex interplay of environmental and biogenetic factors may contribute to PSB. Environmental factors [12] involved may include adverse childhood experiences, family and peer environments, sociocultural factors, and early exposure to pornography [13]. Increasing rates of PSB may be tied to increased sexualized content in the media, including on social media [14], as well as the increase in the breadth and ease of access to pornographic materials [15]. Meanwhile, neurobiological studies examining hypersexuality by looking at the reward system in the brain suggest that alterations in the reward circuit may play a prominent role in its emergence [16]. The reinforcing effects of sex may be altered by dopaminergic and other neurotransmitter-related gene polymorphisms [17], with some evidence of this being observed in candidate gene analyses [18, 19]. Another approach has been looking at neuroendocrine and neurochemical systems and hormones involved in sexual behaviours, finding alterations in oxytocin levels [20] and suggesting the need for research into the role of the hypothalamus-pituitary-adrenal axis, the hypothalamus-pituitary-gonadal axis, and the oxytocinergic systems; genetic variation in such may play a role [21].

Research into the genetic epidemiology of PSB remains relatively sparse due to factors such as the diagnostic reasons listed above, as well as stigma, shame and sociocultural attitudes, but evidence from related conditions and phenotypes supports a genetic basis for both environmental susceptibility and shaping environmental selection. For example, a family study observed that individuals with hypersexuality are likely to have parents with similar conditions, and family members with at least one addiction disorder [22]. Twin studies of related phenotypes (paraphilia and sexual coercive behaviour [23], problematic masturbatory behaviour [24], risky sexual behaviour [25], number of sexual partners[26, 27] and age at first sexual intercourse (AFS) [12, 28]) demonstrate significant heritable components, with heritability ranging from ∼0.30 to 0.80 depending on the cohort and phenotype. Twin studies of addictive disorders, which may share genetic components with PSB, estimate that the heritability of gambling and substance use disorders ranges from *h^2^*∼0.39-0.72 [26], while impulsivity (*h^2^*∼0.50)[29, 30], impulse control disorders (*h^2^*∼0.57) [31], personality disorders (*h^2^*∼0.30-0.60) [32] and ADHD(*h^2^∼*0.74) [33] are also thought to have significant genetic components. Indeed, a twin study observed that risky sexual behaviour was genetically correlated with impulsivity (r=0.27) and extraversion (r=0.24) [34]. Relatedly, delay discounting, the preference of small immediate rewards over delayed, larger rewards, has also been shown to have a significant heritability (*h^2^∼*0.35-0.62) [35, 36]. The idea of a reward deficiency syndrome (RDS) resulting from a breakdown of the reward related neurotransmission due to genetic and epigenetic influences [17, 37, 38], and which encompasses many health disorders including addictions, compulsions and impulsive behaviours, has been proposed to be an underlying mechanism of PSB [9, 18, 39]. Overall, multiple pathways to PSB appear plausible.

Genome-Wide Association Studies (GWAS) allow the exploration of associations between single nucleotide polymorphisms (SNPs) across the whole genome and traits of interest. Through GWAS, it has been shown that the genetic architectures of disorders of addiction [40], personality [41] and behaviour [41, 42] have significant polygenic components. Several GWAS have touched on ideas related to PSB, but there is no prior GWAS of PSB. A GWAS using data from the UK Biobank found 38 variants associated with AFS, with some of these loci being associated with behavioural and reproductive traits [43]. A series of GWAS on risky and externalizing behaviours (including AFS, number of lifetime sexual partners, and substance use) have identified over 500 associated genomic loci [43–46]. Another GWAS identified 4 ancestry and sex-specific genome-wide significant loci for an interaction between alcohol dependence and risky sexual behaviour [47]. Delay discounting was observed to have 11 risk loci and a transdiagnostic genetic component over mental and physical health [48].

Here, we report the first case-control GWAS of PSB. With the knowledge that the sample size available is limited to detect significant markers, we aimed to detect genes and gene-sets with potential involvement in PSB, and using polygenic risk scores (PRS), we explored genetic overlap with psychiatric disorders, and relevant personality traits and behavioural phenotypes.

## PARTICIPANTS AND METHODS

### Participants

#### Patients

Individuals receiving inpatient and outpatient treatment for PSB at 8 addiction treatment facilities in the United States (n=512) were recruited. Of these, n=448 provided DNA samples.

#### Comparison Participants

Full details of the recruitment of the comparison sample have been described elsewhere [9]. Briefly, the Office of the Registrar at the University of Alberta invited 68,846 eligible students (undergraduate and graduate) and postdoctoral fellows by email (all individuals registered 2016–2019) to participate in the study. Interested individuals emailed the study team and were subsequently sent an emailed invitation to review the study details and complete an online consent form, followed by screening measures.

Inclusion criteria were: ≥18 years of age, undergraduates, graduates, postdoctoral fellows, and recently convocated students registered in at least 1 course in the preceding year, excluding those who had withdrawn after registration without any reason to return in the subsequent term or be on campus (e.g., suspension), and the ability to answer in English. Of n=3341 participants, n=1742 provided saliva samples for DNA.

#### Ethics

All participants provided written informed consent. The Quorum Review Institutional Review Board (Seattle, Washington; Protocol Number: 2016-001) and the University of Alberta Research Ethics Board (Protocol: Pro00066552) approved the study.

### Assessments

#### PSB

The 20-item Sexual Addiction Screening Test - SAST-R Core was used to screen for PSB in comparison patients, as previously described [9]. It measures 4 addictive dimensions of PSB: relationship disturbance, loss of control despite problems, preoccupation with sex, and affect disturbance. After recruitment, it was observed that some university participants met criteria for PSB (SAST-R Core population level threshold score ≥6) [9]. All patients were considered to have PSB.

#### Reward Deficiency Syndrome

An indicator for RDS was derived by combining measures screening for several relevant conditions, including nicotine dependence (Fagerström Test for Nicotine Dependence [FTND]), pathological gambling (DSM-5 Pathological Gambling Diagnostic Form [DPGDF]), personality disorders (8-item Standardized Assessment of Personality-Abbreviated Scale as a Self-Administered Screening Test [SA-SAPAS]), ADHD (Adult ADHD Self-Report Scale [ASRS] version 1.1), compulsive buying (Richmond Compulsive Buying Scale [RCBS]), and internet addiction (Internet Addiction Test [IAT]), as previously described [9]; giving a binary variable (yes if respondent screened positive for any assessment, otherwise no).

#### Sample Collection and Genotyping

Patients provided samples using OG-500 kits (Oragene, DNA Genotek, Kanata, Canada). For comparison participants, DNA was primarily sampled using buccal swabs (4 swabs per person) or from saliva additionally collected from participants from whom DNA yield was low. DNA was extracted from the buccal swabs using the Xtreme DNA Isolation Kit (Isohelix, D-Mark Biosciences, Toronto, Canada), and from saliva according to the manufacturer’s recommendations.

Genotyping was performed using the Infinium Global Screening Array-24 v3.0 BeadChip with the Psych Booster add-on (Illumina Inc, San Diego CA, USA), with 698,585 variants. Genotyping and quality control (QC) were performed according to standard procedures. Genetic data were available from n=2132 individuals (n=448 patients and n=1684 comparison participants [comparison PSB n=196, controls n=1488]).

#### Data Analysis and Quality Control

GenomeStudio 2.0.5 (Illumina, Inc.) was used for analyzing array data. Data quality control and association analyses were performed using Plink 2.0 [49, 50] using standard methods for SNP and individual filtering. Briefly, the following parameters were used: subject missingness >0.1; autosomal heterozygosity deviation (|Fhet|> 0.2), SNP missingness>0.02; case/control difference in SNP missingness (0.02); Hardy-Weinberg Equilibrium (HWE) <1e-6; Minor Allele Frequency (MAF) 0.01. 6 samples were removed due to sex mismatch. Analyses were limited to autosomal SNPs. Relatedness testing was performed using the KING algorithm, with suspected inbreeding n=1, and one of the pair of cryptically related subjects (cut-off, 0.088, second-degree relation n=61 removed. The majority of the sample (>60%) comprised individuals of European ancestry (EUR), but also included individuals of Asian (East, South, Southeast, West), Arab, Black, Indigenous, Latin American, and Pacific Islander ancestry. We analyzed the data separately as EUR only and All Ancestry (AllAnc) datasets as detailed below.

Population stratification was dealt with using Principal Component Analysis (PCA). Two rounds of PCA using 10 principal components (PCs) were conducted, using an LD-filtered SNP set (MAF:0.01, SNP missingness geno:0.02, Individual Missingness: 0.02, HWE 1e-6, LD pruning: indep-pairwise 200 50 0.1) removing genetic outliers (greater than 6SD, EUR: n=22, AllAnc: n=54). 10 PCs were generated from the filtered set and used as covariates in subsequent analyses. This was done separately for the EUR and AllAnc datasets.

After QC steps and filtering missing phenotypic data, the datasets comprised EUR (total n=1177; PSB patient n=337, comparison PSB n=80, controls n=760), AllAnc (total n=1947; PSB patient n=379, comparison PSB n=176, control n=1339) individuals with 473,244 variants. Downstream analyses were performed separately in both EUR and AllAnc datasets.

#### Association Analysis

Two logistic regression models were used to test for single marker associations with PSB. The first model (“All-PSB Model”) defined case/control status using PSB patients & PSB comparison participants vs. non-PSB comparison participants. The second model defined case/control status (“Patient-PSB Model”) by comparing PSB patients vs. comparison participants without PSB. A third regression model was used with the RDS phenotype (Yes vs. No).

#### Gene-based, gene-set, tissue expression analysis and pathway analysis tests

GWAS data were used for gene-based, gene-set, and tissue expression tests, as well as gene mapping, as implemented in SNP2GENE of FUMA v1.52 [51] accessed 2025-09-29), which incorporates MAGMA v1.08[48]. Gene-based tests were performed using the summary statistics of the models for a total of 17,746 protein coding genes based on the physical position of SNPs. SNPs were included using boundaries of 10 kilobases (kb) upstream and downstream of the genes. Gene-sets were tested by analyzing 17,009 (MSigDB v7.0) gene-sets. Bonferroni correction was applied for both gene-based and gene-set tests, correcting for the number of respective tests (gene-based p<0.05÷17,746=2.8618×10^-6^; gene-set p<0.05/17,009=2.94×10^-6^). MAGMA gene expression analysis using GTEx v8 54 tissue types and 30 general tissue types was performed to map genetic variants to potentially relevant tissue-specific gene expression, identifying which tissues and biological pathways might be most relevant to PSB.

#### Polygenic Risk Scoring

Polygenic risk scores (PRS) were calculated for each individual using PRSice2[52] based on summary statistics for psychiatric disorders, personality traits and behaviours which have been suggested to be associated with PSB (**Table 1**). All summary statistics used were in populations of European ancestry. PRS were calculated by summing allele counts of SNPs weighted by GWAS effect sizes. The extended major histocompatibility complex region (Chr 6: 26-33MB) was excluded due to its LD structure. Effect sizes were standardized. PRS were calculated for various p-value thresholds (5x10^-8^, 1x10^-6^, 1x10^-4^, 0.001, 0.01, 0.05, 0.1, 0.2, 0.5, 1). Association of the most informative PRS with case-control status was calculated for each model (All-PSB vs. Controls, Patients vs Controls, RDS (yes/no)) including genetic 10 PCs as covariates, and only European ancestry samples were used. Bonferroni corrections for multiple testing were used (p=0.05/84 [3 comparisons x 28 phenotypes] =0.000595).

**Table 1.** Summary statistics used in Analyses. Age at first Sex and Number of Lifetime Sexual Partners are incorporated in Externalizing summary statistics. European ancestry summary statistics, without 23 and me samples included, were used in all cases. BIS: Barratt Impulsiveness Scale. UPPS-P: UPPS-P Impulsive Behavior Scale.

| Phenotype | N | Cases | Controls | Reference |
| --- | --- | --- | --- | --- |
| *Psychiatric Disorders* |  |  |  |  |
| Anxiety Disorders (Case Control) |  | 5,761 | 11,765 | Otowa et al., 2016[83] |
| Attention Deficit Hyperactivity Disorder |  | 38,691 | 186,843 | Demontis et al., 2023 [84] |
| Borderline Personality Disorder |  | 12,339 | 1,041,717 | Streit et al., 2025 [85] |
| Bipolar Disorder |  | 59,287 | 781,022 | O'Connell et al., 2025 [86] |
| Major Depression |  | 412,305 | 1,588,397 | MDD Working Group et al., 2025 [87] |
| Obsessive-Compulsive Symptoms | 33,943 |  |  | Strom et al., 2024 [58] |
| Obsessive-Compulsive Disorder |  | 53,660 | 2,044,417 | Strom et al., 2025 [59] |
| Posttraumatic Stress Disorder | 641,533 |  |  | Stein et al. 2021 [88] |
| Substance Use Disorders | 647,703 |  |  | Hatoum et al., 2023 [40] |
| *Behaviours* |  |  |  |  |
| Age at First Sexual Intercourse | 387,338 |  |  | Mills et al., 2021 [45] |
| Broad Antisocial Behaviour | 85,359 |  |  | Tielbeek et al., 2022 [89] |
| Drug Experimentation | 23,111 |  |  | Sanchez-Roige et al. 2019 [73] |
| Externalizing | 1,045,957 |  |  | Karlsson Linnér et al., 2022 [44] |
| Number of Lifetime Sexual Partners | 378,882 |  |  | ieu open GWAS project (ukb-b-4256) [90] |
| *Big Five Personality* |  |  |  | Gupta et al., 2024 [41] |
| Openness | 237,390 |  |  |  |
| Conscientiousness | 252,253 |  |  |  |
| Extraversion | 298,772 |  |  |  |
| Agreeableness | 252,749 |  |  |  |
| Neuroticism | 682,688 |  |  |  |
| *Impulsivity* |  |  |  | Sanchez-Roige et al. 2019 [73] |
| BIS Attentional | 22,385 |  |  |  |
| BIS Motor | 22,306 |  |  |  |
| BIS Nonplanning | 22,291 |  |  |  |
| BIS Total Score | 21,972 |  |  |  |
| UPPS-P Negative Urgency | 23,344 |  |  |  |
| UPPS-P Perseverance | 23,437 |  |  |  |
| UPPS-P Positive Urgency | 23,284 |  |  |  |
| UPPS-P Premeditation | 23,321 |  |  |  |
| UPPS-P Sensation Seeking | 23,292 |  |  |  |

### LD Score Regression: Proxy Genetic Correlations and Heritability

While the primary sample did not reach the recommended size (n∼5000) [53] to perform genetic correlations; as a proxy, to assess the genetic association structure, we calculated pairwise correlations using a subset of the summary statistics used to perform PRS calculations using LD Score Regression (LDSC) [54]. With size limitations in mind, as an exploratory step, using univariate LDSC, we estimated SNP-heritability using the results from the European ancestry All-PSB, Patient-PSB and RDS models.

## RESULTS

### Association Analyses and FUMA

#### European Ancestry

No significant genome-wide associations were observed with single variants in all models. Gene-based tests and MAGMA gene-set analysis and Tissue Expression analyses did not find significant associations after multiple-testing correction. Below, we report suggestive associations and top hits; details can be found in the Supplementary Material (**Tables S1-3, Figure S1abc**).

In the All-PSB vs. Controls model, the top hits comprised two in close proximity on Chr 11 (one suggestive, rs10770107, p=9.05x10^-6^; rs6484218, p=1.44x10*^-5^*) and one on Chr 9 (rs10118434, p=1.46x10^-5^) (**Table S1**). In the Patient model, the top hits included the same variant as the All-PSB model on Chr 9 (suggestive, rs10118434, p=2.22x10^-6^) **(Table S2)**. In the RDS model, top hits included two suggestive variants on Chr 8 (rs75778290, p=2.16x10^-6^) and Chr 4(rs11723502, p=3.64x10^-6^) (**Table S3**).

#### All Ancestries

No significant genome-wide associations were observed with single variants in all models. Below, we report suggestive associations and genomic loci identified by FUMA; details can be found in the Supplementary Material (**Tables S4-6, Figure S1def**)

#### All-PSB vs. Controls

Three genomic loci showed suggestive associations (< 1x10^-5^) in the single variant analyses, two on chromosome 11 (rs6484218, p=4.69x10^-7^; rs11214998, p=9.37x10^-6^) and one on chromosome 20 (rs6108761, p=9.22x10^-6^) (**Table S4ab**). These mapped to 11 genes; details are shown in **Table S4c**. The gene-based test did not yield any genome-wide significant genes (p=0.05/17748=2.82x10^-6^).

In the Gene-Set analysis, two gene-sets were significantly associated after Bonferonni correction; GOMF_LEUCINE_ZIPPER_DOMAIN_BINDING (n genes=10, p_Bon_=0.00021) and GOMF_LRR_DOMAIN_BINDING (n genes=15, p_Bon_=0.0040). (**Table S4d)**

In the tissue type analyses, no tissues were significantly associated after multiple testing correction (54 types, 0.05/54 = 0.0009, -log_10_ = 3.03; 30 types p = 0.05/54 = 0.00167, - log_10_=2.78). Several tissues were nominally (p < 0.05) associated: using 54 types, adrenal gland (p=0.006), ovary (p=0.01), cervix_ectocervix (p =0.029); using 30 types, adrenal gland (p=0.027) and ovary (p=0.039). **(Figure S2ab)**

#### Patients vs. Controls

Three genomic loci showed suggestive associations in the Patient model. One on Chr7 (rs10234084, p=2.29785x10^-6^ and two on Chr 11 (rs6484218, p=2.90x10^-6^; rs2276410, p=3.53x10^-6^) **(Table S5ab)**, mapping to 5 genes (**Table S5c)**.

In the gene-set analysis, two gene-sets were significantly associated after Bonferonni correction; GOMF_LEUCINE_ZIPPER_DOMAIN_BINDING (n genes=10, pBon=0.0032) and GOMF_LRR_DOMAIN_BINDING (n genes=15, pBon=0.0098) (**Table S5d)**.

In the tissue type analyses, no tissues were significantly associated after multiple testing correction (thresholds as above). Several tissues were nominally associated: using 54 types, uterus (p=0.019), ovary (p=0.019), coronary artery (p=0.027), and adrenal Gland (p=0.027), spleen (p=0.032), cervix-endocervix (p=0.036); using 30 types: Ovary (p=0.045), Uterus (p=0.046), and Spleen (p=0.049) **(Figure S2cd)**.

#### RDS

One genomic locus was identified showing suggestive association with RDS, rs7655156 on Chr 4 (p=8.59x10^-6^), mapping to 2 genes (**Table S6abc**). The single gene analysis did not identify any significant or suggestive associations. The gene-set analysis found one gene-set, GOMF_STRUCTURAL_CONSTITUENT_OF_TOOTH_ENAMEL, to be significantly associated with RDS (p_bon_=0.00059) (**Table S6d**).

In the tissue type analyses, no tissues were significantly associated at corrected thresholds. Nominal associations were observed using 54 types: stomach (p=0.030); using 30 types, stomach (p=0.011) and small intestine (p=0.032) **(Figure S2ef)**.

### Polygenic Risk Scores

PRS for the various phenotypes were significantly associated with PSB status (both All-PSB and Patient-PSB models), and associations had both similarities and differences with RDS (**Fig 1**). Patient-PSB was more strongly associated with the majority of PRS compared to All-PSB and RSD. Detailed results of PRS analyses are shown in **Tables S7-9**.

**Figure 1.**
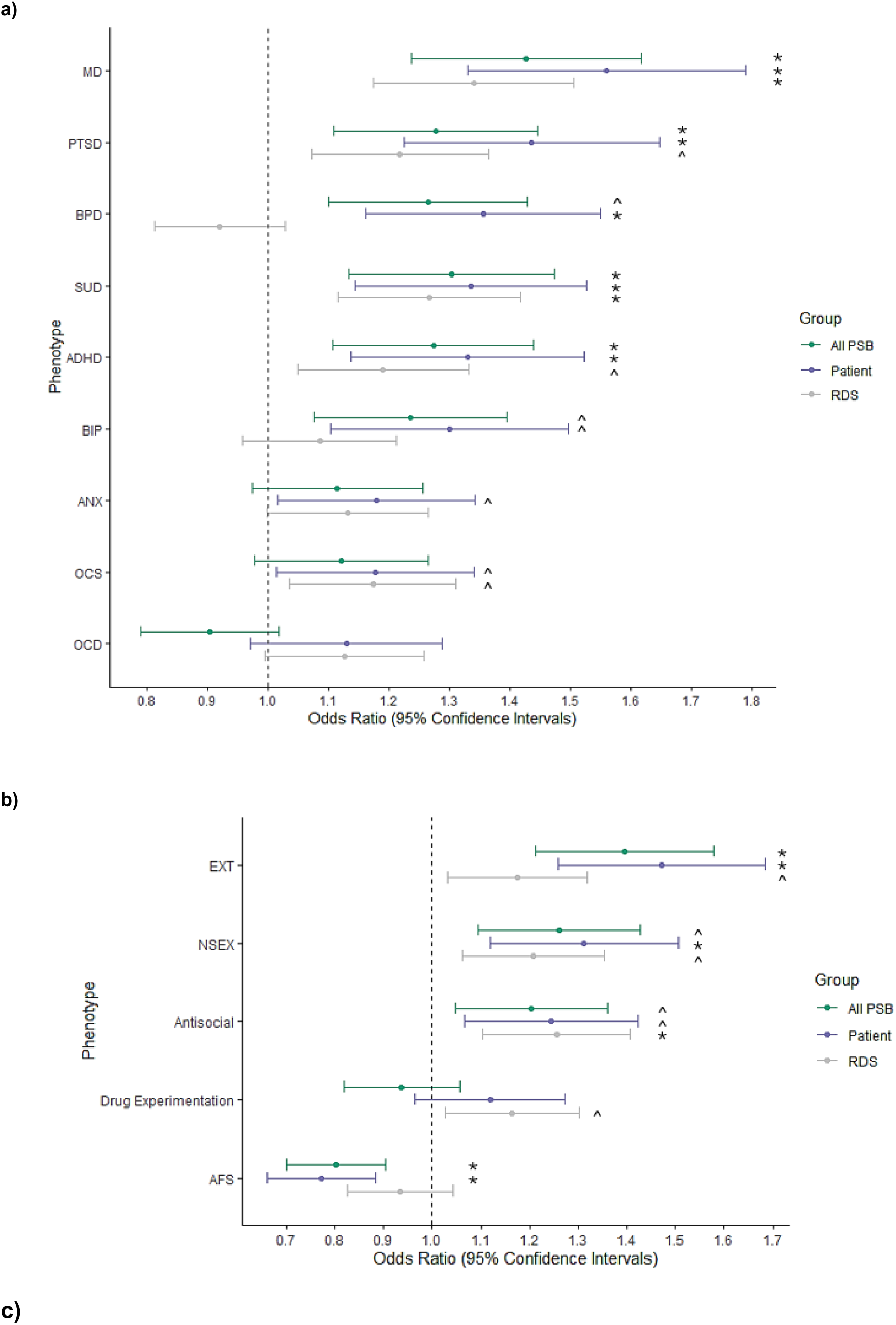
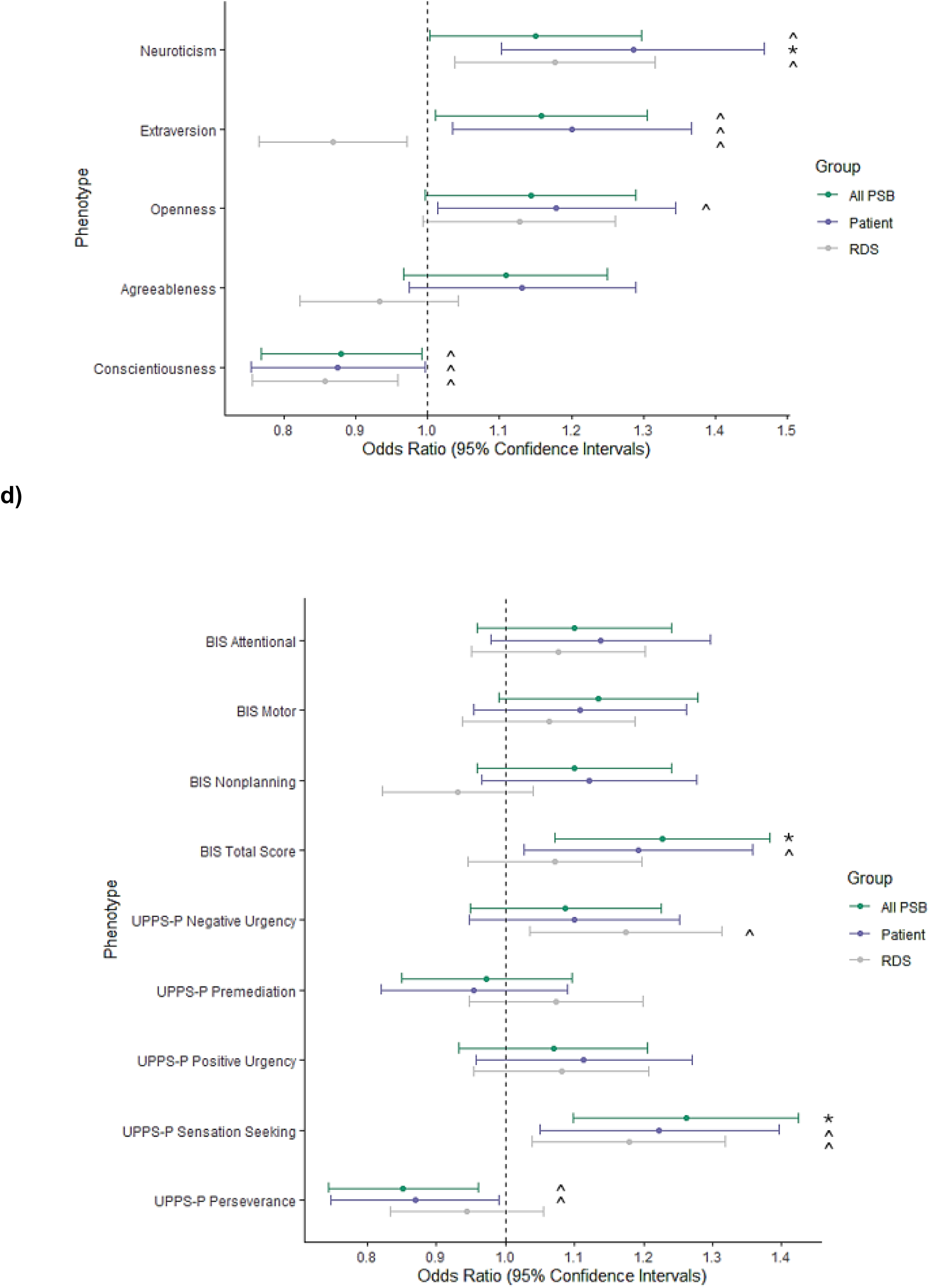
Polygenic risk scores for a) psychiatric conditions, b) behaviours and c) BIG5 personality traits d) impulsiveness personality traits and associations with PSB and RDS. AFS: Age at First Sexual Intercourse, Antisocial: Broad Antisocial Behaviour, ANX: Anxiety, ADHD: Attention Deficit Hyperactivity Disorder, BIS: Barratt Impulsiveness scale. BPD: Borderline Personality Disorder, Extra: Extraversion, EXT: Externalising, MD: Major Depression, NSEX: Number of Lifetime Sexual Partners, OCD: Obsessive Compulsive Disorder, OCS: Obsessive Compulsive Symptoms, PTSD:Posttraumatic Stress Disorder, SUD: Substance Use Disorders, Open: Openness, Consc: Conscientiousness, Extra: Extraversion, Agree: Agreeableness, Neuro: Neuroticism. UPPS-P: UPPS-P Impulsive Behavior Scale. *significant after Bonferroni correction (p<0.05/84=0.000595) ^ p<0.05

PRS generated using psychiatric disorder summary statistics (**Fig. 1a**) were positively significantly associated with Patient-PSB status (major depression [MD], post-traumatic stress disorder [PTSD], borderline personality disorder [BPD], substance use disorder [SUD], ADHD, all p<0.000595), with bipolar disorder (BIP) and anxiety disorders (ANX) nominally associated (p<0.05). While PRS for Obsessive Compulsive Symptoms (OCS) were nominally associated with Patient-PSB (p<0.05), PRS for Obsessive Compulsive Disorder (OCD) were not. All-PSB was significantly associated with MD-, PTSD-, SUD- and ADHD-PRS (all p<0.000595) and nominally with BPD- and BIP-PRS (p<0.05). RDS was significantly associated with MD- and SUD-PRS (both p<0.000595), and nominally with PTSD-, ADHD- and OCS-PRS (all p<0.05).

Examining PRS for behavioural phenotypes (**Fig. 1b**) found that PRS for externalising behaviours were significantly associated with both PSB definitions (p<0.000595), and nominally with RDS (p<0.05). PRS for AFS was significantly associated with both PSB definitions in the negative direction (both p< 0.000595); younger AFS was associated with All-PSB and Patient PSB. NSEX-PRS were significantly associated with Patient PSB (p<0.000595), with All-PSB and RDS showing nominal associations (p<0.05). Broad antisocial behaviours were associated with RDS (p<0.000595) and both PSB definitions (both p<0.05). Drug Experimentation-PRS were associated with RDS at the nominal level (p=0.017), but not both definitions of PSB.

For the Big 5 personality traits (**Fig. 1c**), neuroticism-PRS positively were associated with Patient PSB (p<0.000595), and All-PSB and RDS at the nominal level (p<0.05). Nominal associations were observed between conscientiousness-PRS and both PSB definitions and RDS, all in the negative direction (p<0.05). At the nominal level, extraversion-PRS were positively associated with All-PSB and Patient PSB, but negatively with RDS. Openness-PRS were positively associated with only Patient PSB (p<0.05). Agreeableness-PRS were not associated with either PSB definition of RDS.

Looking at impulsivity personality trait-related PRS (**Fig. 1d**), Barratt Impulsiveness Scale (BIS) total score-PRS were associated with both PSB definitions (All-PSB, p< 0.000595; Patient PSB, p<0.05), but not RDS; no BIS subscales were associated with either PSB or RDS. On the UPPS-P, sensation seeking-PRS were associated with both definitions of PSB (All-PSB, p<0.000595; Patient PSB, p<0.05) and RDS (p<0.05), while (lack of) perseverance-PRS were negatively associated with All-PSB and Patient PSB at the nominal level (p<0.05).

### LD Score Regression

#### Genetic Correlations

Significant correlations were observed between the phenotypes used to perform PRS analyses in the 3 models (**Figure S3**). Correlations were observed between the psychiatric disorders, in line with PRS results. Significant correlations were also observed between psychiatric disorders and externalising behaviours, including AFS (negative correlation) and NSEX; no correlations with externalising behaviours were observed with OCD or OCS. Of the personality traits, Neuroticism showed the strongest correlations with psychiatric disorders.

#### SNP Heritability

The estimated SNP heritability on the observed scale for the All-PSB and Patient-PSB models were similar and suggested a polygenic signal (All-PSB: h^2^: 0.6252 (SE=0.5574) (Intercept:1.0049 (SE=0.0097), mean Chi^2^: 1.0164, ratio: 0.2992 (SE=0.5901); Patient: h^2^: 0.5653 (0.5982), intercept: 1.0067 (0.0098), mean Chi^2^: 1.0162, ratio: 0.4117 (0.6038)), while the estimate was lower for RDS (h^2^: 0.1758 (SE=0.5416), intercept: 0.9982 (SE=0.0098), mean Chi^2^:1.0015, ratio:<0), with calculations showing high standard error, reflecting the limited sample size.

## DISCUSSION

The present study is the first clinical case-control GWAS of PSB. While no genome-wide significant associations were found for single variants or genes, several suggestive candidates were raised. PRS for multiple psychiatric disorders, behaviours and personality traits were associated with PSB status, suggesting shared genetic underpinnings, and shedding new light onto the discussion of the classification of PSB.

One variant (rs6484218), suggestive across both the European and All Ancestry PSB analyses, was the top hit in an earlier cross-disorder GWAS of schizophrenia, depression and bipolar disorder [55]. The locus maps to the *AMPD3* gene, which encodes an enzyme that breaks down AMP and plays a crucial role in metabolic processes and inflammatory response modulation (<u>nih.gov/gene</u>). The variant also lies near the adrenomedullin gene (ADM), which is widely expressed in the brain; preclinical research suggests that a lack of CNS adrenomedullin can result in increased anxiety [56]. Due to limited sample size, these results should be viewed as hypothesis generating, nevertheless this finding aligns with the idea of shared genetic vulnerabilities to PSB and other psychiatric disorders.

PRS for psychiatric disorders were significantly associated with PSB status, more strongly for Patient-PSB than All-PSB. MD-PRS having the strongest association is consistent with observed comorbidity; the relationship appears to be bi-directional, with guilt and shame resulting from PSB worsening depressive symptoms, and depression leading to PSB when individuals seek a fix for negative emotions, creating a compensatory cyclical pattern [57]. Both may involve deficiencies in emotional regulation and the reward system; these ideas are supported by the positive association of RDS status with MD-PRS.

SUD-PRS were associated with both definitions of PSB, and RDS to a similar level; this appears consistent with the idea that the ‘addiction’ component of PSB involves the reward system [37]. Comorbid substance use is often observed in PSB, and shared genetics involving reward deficiency may explain the relationship with other psychiatric disorders (e.g., PTSD, MD) in which addiction is also comorbid; a lack of reward sensitivity may play a causal role.

While obsessive compulsive symptom PRS (generated from a GWAS in the general population [58]) were associated with both Patient-PSB and RDS at the nominal level, PRS from the case-control GWAS of OCD [59] did not, suggesting limited overlap between PSB and OCD. It may be that the more severe forms PSB (i.e., our patient sample) do not have a strong overlap with factors such as obsessive fear/anxiety leading to compulsive behaviour; the behaviour in PSB may be driven by other factors. Given the complexity of these underlying mechanisms and heterogeneity of PSB [60] and our prior data [36], our results suggest that for the majority of PSB patients, an addiction may be a better description than an impulse control disorder.

Taken together, these results may inform the scientific discourse on the classification of PSB. CSBD is currently classified under Impulse Control Disorders (Code 6C72), but not as a behavioral addiction in the ICD-11 [6]. The manifestation of PSB in the present sample has genetic overlap with addiction, and, to a lesser degree, with compulsion; research in larger, more comprehensive samples is needed to further characterise this relationship.

PRS for BPD, ADHD and broad antisocial behaviours, which are strongly genetically correlated, were positively associated with both definitions of PSB, suggesting that genetic vulnerability to poor self-regulation and impulsivity, as well as affective instability and emotional reactivity associated with higher sensitivity to stress and external stimuli, may be common across these phenotypes. Of these, only the BPD-PRS was not associated with RDS, which may be consistent with the profile of BPD. The relationship between ADHD and hypersexual symptomatology may be mediated by depressive feelings [61] and the overlap of genetic mechanisms may be similar to that with MD; the presence of a neurodevelopmental disorder is a known risk factor for PSB [62]. Accordingly, difficulties with sustained goal-directed engagement (establishing/maintaining stable intimate relationships requires ongoing commitment, persistence and investment) as observed in ADHD may suggest a subtype of PSB characterized by dysregulated relational pursuit and attachment rather than by reward-driven sexual behaviour [63]. The association of broad antisocial behaviour-PRS with both PSB and RDS may support the hypo-reactivity model of reward thought to drive these behaviours; dysfunction in brain regions, specifically lower striatal reactivity to reward cues and lower prefrontal-regulatory recruitment during reward and loss anticipation may collectively underlie the persistence of delinquent behaviours, such as disregard for social norms and for the wellbeing of others despite negative consequences [64].

The Externalizing-PRS results offer insight into the genetics underlying PSB; Karlson-Linner et al. [44] modeled ADHD, problematic alcohol use, cannabis initiation, smoking initiation, AFS, NSEX and general risk tolerance by a single underlying genetic factor representing the shared genetic liability to these behaviors and a core feature of impulsivity. In the present study, EXT-PRS was significantly associated with PSB, as were the NSEX- and AFS-PRS (both incorporated in EXT summary statistics) [44]. RDS was associated to a lesser extent than PSB with EXT-PRS, and not with AFS, but NSEX was associated with RDS to a similar level as PSB. This may reflect the different components covered by the two phenotypes; AFS may reflect genetics involved in initiation but may also be related to childhood trauma and environmental sex as triggers, while also being linked to maladaptive future patterns of sexual behaviour [65, 66]. NSEX may be an indicator of magnitude, maintenance and continuation of behaviours, which may qualitatively be connected to promiscuity and novelty seeking. The pattern of genetic correlations of EXT, NSEX and AFS with psychiatric disorders shows positive (NSEX, EXT) and negative (AFS) correlations with all of the tested disorders, but none with OCD, which may inform the debate about the nature of PSB. Meanwhile, we observe significant genetic correlations between OCD and the other disorders, suggesting specific differences in genetic overlap.

Of the Big 5 personality traits, Neuroticism-PRS was most strongly associated with (Patient) PSB, mirroring findings of strong genetic correlations between neuroticism and psychiatric disorders in the literature [67]. Openness being associated with patient (severe) PSB but not milder PSB or RDS may be related to a novelty-driven or boundary-testing component confined to the more severe PSB. Conscientiousness being negatively associated with PSB/RDS and agreeableness showing no association appear consistent with those traits.

Impulsive personality traits have been studied in respect to risky sexual behaviour [68–70], which overlaps with but is not equivalent to PSB. Impulsive personality traits are complex and have been conceptualized with several different facets; we found similarities and differences in the associations of the respective PRS with PSB and RDS, which aligns with the idea that various aspects of impulsivity have differential associations with PSB [71]. Total BIS score was associated with both definitions of PSB, consistent with observations from the phenotypic literature implicating general impulsivity in PSB [57, 72]; no association was observed with RDS. The positive association of Sensation Seeking with both PSB definitions and RDS suggests a shared mechanism; the impulsivity GWAS [73] implicated the *CADM2* locus, which has also been associated with risk taking propensity/behaviour, alcohol consumption [74], and NSEX and number of children [46]. It [73] also identified associations in the *CADM2* locus with drug experimentation; in the present study, drug experimentation-PRS were associated with RDS, but not PSB, suggesting an independent mechanism. Negative urgency-PRS were associated with RDS, but not PSB; the genetics underlying tendency to experience strong impulses under conditions of negative affect may more strongly drive RDS related behaviours than PSB. Interestingly, the (lack of) perseverance-PRS – a failure to tolerate boredom or to remain focused despite distraction, leading to difficulty in following through with tasks – were negatively associated with both PSB definitions (but not RDS); this may reflect the need to keep focus to carry out actions related to PSB, such as planning and navigating potential barriers. However, (lack of) premeditation-PRS (acting without prior thought/planning and inability to think through consequences) were not associated with PSB or RDS, which may be consistent with fewer findings and weak/mixed results about the connection between perseverance and premeditation with risky sexual behaviour [68, 69].

Gene-set analyses in both PSB models found enrichment in the GOMF_LEUCINE_ZIPPER_DOMAIN_BINDING and GOMF_LRR (Leucine Rich Repeats)_DOMAIN_BINDING gene-sets; Leucine zipper motifs affect gene transcription, and like LRR domains, which mediate protein-protein interactions, and are relevant to immune response [75, 76]. There is increasing recognition of the complex relationship between immune function and mental health [77]. While it is unclear what the role of the significant gene-set for RDS (GOMF_STRUCTURAL_CONSTITUENT_OF_TOOTH_ENAMEL) is, it is worth noting that mental and oral health are linked, and people with severe mental illness have higher rates of dental issues [78, 79].

The present study had several limitations. As the first case-control PSB study, the sample size is small, and the definitions for PSB/RDS used may not generalize to other samples. Further exploration in larger cohorts and independent cohorts remains. While we included all ancestry in our analyses, our primary analyses focused on individuals of European ancestry, given the makeup of the sample and available summary statistics. This highlights the need for future work in samples of diverse ancestry, given not only genetic and biological but also sociocultural factors potentially involved. The demographics of the patient and comparison samples could not be matched (e.g., age and sex); the comparison sample comprised students and postdoctoral fellows recruited from a university, leaving a possibility of ascertainment bias; the frequency of PSB observed was comparable to general population figures [36]. Our sample reflects the known observed higher prevalence of PSB in males, greater representation of women is needed in future work. Our measure of RDS was a composite score based on an aggregation of multiple scales; at the time of data collection, research on RDS had not yet arrived at measures such as the RDS Questionnaire [80]; subsequent research should leverage up-to-date assessment tools. The SNP heritability estimates are preliminary and will need confirmation in larger samples. Finally, we note that PRS analyses do not give point estimates of effect magnitude *per se*, but are also driven by the power of the discovery GWAS and other factors; significant variation explained by the models was modest (∼1.4-4.5%), as observed in the literature (**Tables S7-S9**).

## CONCLUSION

Genetic factors may predispose to a dysphoric state; the nature of that state being the result of environmental experience, with depression being brought about by losses, and anxiety being brought about by constant exposure to danger [81]. Our results raise the possibility of a similar mechanism involved in the development of PSB; childhood sexual abuse, or early exposure to developmentally inappropriate materials may play the environmental role. PSB appears to have underlying genetics enriched for broad biological processes shared with a wide number of psychiatric illnesses [82] and behaviours. Our treatment sample may capture a subgroup of PSB more strongly associated with certain psychiatric disorders and addiction, underlining that further research into genetic and environmental aetiology is needed to improve understanding, and prospects for intervention and treatment.

## Data Availability

Summaries of the data produced in the present study may be available upon reasonable request to the authors

## Acknowledgements

The work herein was supported by an Alberta Centennial Addiction and Mental Health Research Chair and transitional funding (to KJA), Canada Foundation for Innovation (CFI), John R. Evans Leaders Fund (JELF) grant (32147 - Pharmacogenetic translational biomarker discovery), Alberta Innovation and Advanced Education Small Equipment Grants Program (to KJA), and a research grant and philanthropic support from the American Foundation for Addiction Research (to KJA). A Fulbright-Canada-Palix Foundation grant (to PJC) assisted with his contributions to study design, project management, and collaborative working. SJ was supported by an Alberta Innovates Postdoctoral Recruitment Fellowship.

