## Supplementary Information for "A genome-wide association study of problematic sexual behaviour: genetic overlap with psychiatric, behavioural and personality phenotypes"

**Running Title:** A GWAS of Problematic Sexual Behaviour

**Authors:**
Jerome Clifford Foo, PhD[1,2,3], Shui Jiang, PhD[1], Yaroslav Ilnytskyy, PhD[4], Dongping Li, PhD[4], Xiuying Hu BM, MSc[1], Randolph Arnau, PhD[5], Rick Isenberg, MD[6], Bradley Green, PhD[7], Igor Kovalchuk MD, PhD[4], Josef Frank, PhD[3], Rohit Lodhi MD, PhD[8], Leslie Roper MSc MC[1], David Wishart, PhD[9,10,11,12,13], BASIC[14^], Fabian Streit, PhD [15,16,17], Patrick J Carnes, PhD[18], Katherine Jean Aitchison, BM BCh, PhD, FRCPsych[1,2,19,20*]

**Affiliations:**
[1] Department of Psychiatry, College of Health Sciences, University of Alberta, Edmonton, AB, Canada
[2] Neuroscience and Mental Health Institute, University of Alberta, Edmonton, AB, Canada
[3] Department of Genetic Epidemiology in Psychiatry, Central Institute of Mental Health, Medical Faculty Mannheim, University of Heidelberg, Mannheim, Germany
[4] Biological Sciences, Faculty of Arts & Science, University of Lethbridge, Lethbridge, AB, Canada
[5] School of Psychology, University of Southern Mississippi, Hattiesburg, MS, USA
[6] Psychological Counselling Services, Ltd., Scottsdale, AZ, USA
[7] Department of Psychology and Counseling, University of Texas at Tyler, Tyler, TX, USA
[8] Department of Psychiatry, Schulich School of Medicine and Dentistry, Western University
[9] Department of Biological Sciences, University of Alberta, Edmonton, AB, Canada
[10] The Metabolomics Innovation Centre (TMIC), Edmonton, AB, Canada
[11] Department of Computing Science, University of Alberta, Edmonton, AB, Canada
[12] Department of Laboratory Medicine and Pathology, University of Alberta, Edmonton, AB, Canada
[13] Faculty of Pharmacy and Pharmaceutical Sciences, University of Alberta, Edmonton, AB, Canada
[14] Behavioral Addictions Studies and Insights Consortium*
[15] Hector Institute for Artificial Intelligence in Psychiatry, Central Institute of Mental Health, Medical Faculty Mannheim, Heidelberg University, Mannheim, Germany
[16] Department of Psychiatry and Psychotherapy, Central Institute of Mental Health, Medical Faculty Mannheim, Heidelberg University, Mannheim, Germany
[17] German Center for Mental Health (DZPG), Partner Site Mannheim - Heidelberg - Ulm, Germany
[18] The Meadows, Wickenburg, USA
[19] Department of Medical Genetics, College of Health Sciences, University of Alberta, Edmonton, AB, Canada

[20] Psychiatry Section, Division of Clinical Sciences, Northern Ontario School of Medicine, Thunder Bay, ON, Canada

^Consortium members listed in supplementary material

***Correspondence to:**
Katherine Aitchison, BM BCh, PhD, FRCPsych
Tel number: 780-492-4018

5-020, Katz Group Centre for Pharmacy and Health Research, Edmonton, AB, Canada

**Figure S1. Manhattan plots. a)** All-PSB, European; **b)** Patient PSB, European, **c)** RDS, European; **d)** All-PSB, All Ancestry; **e)** Patient PSB, All Ancestry; **f)** RDS, All Ancestry. Red line indicates genome wide significance threshold: 5 x $10^{-8}$. Blue line indicates suggestive significance threshold: 1 x $10^{-5}$.

**a)**

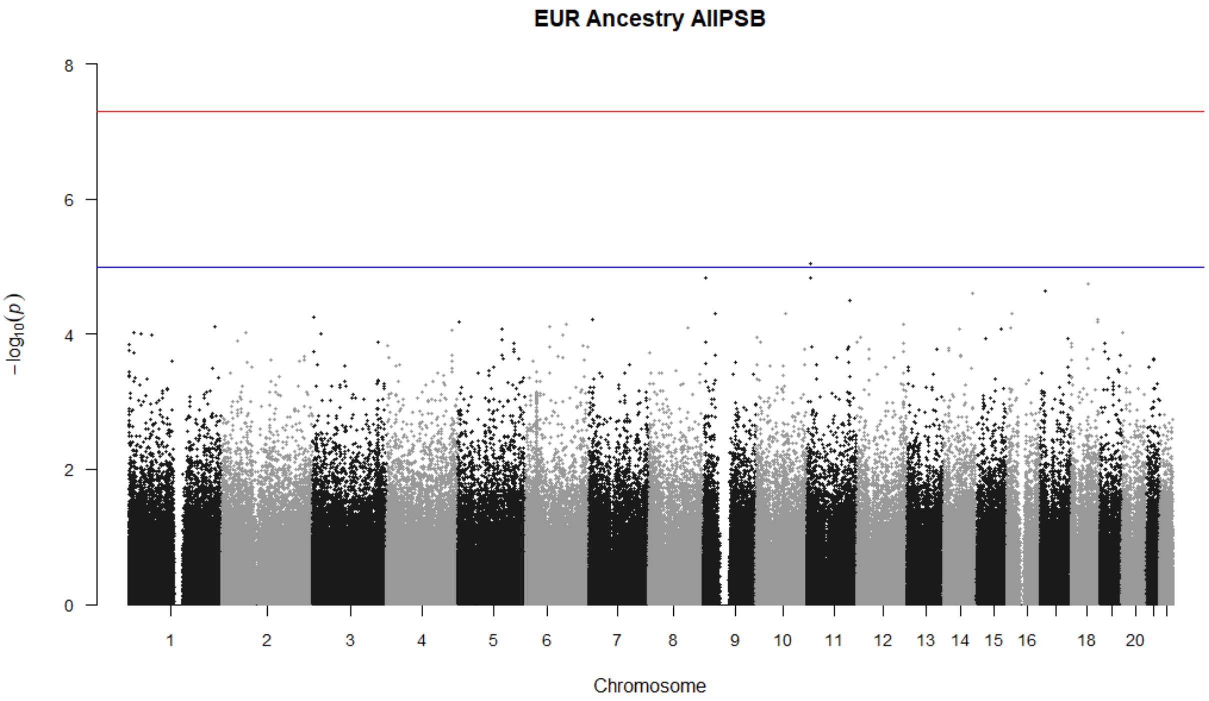

**b)**

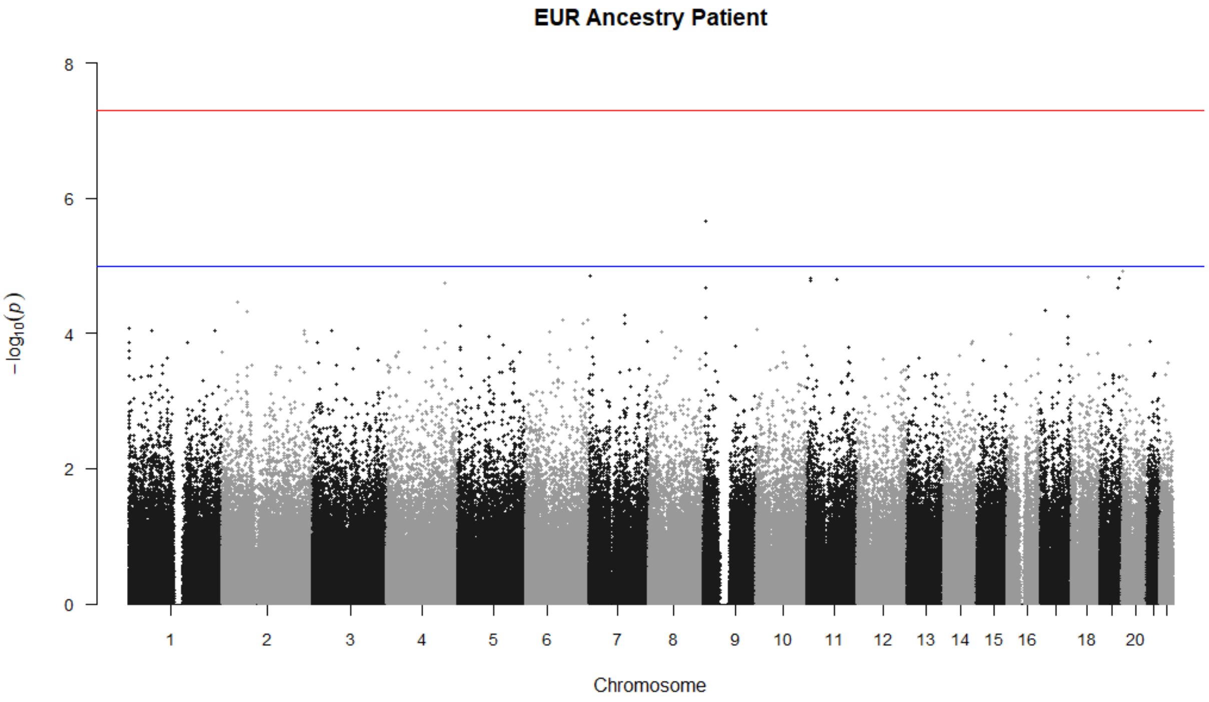

**c)**

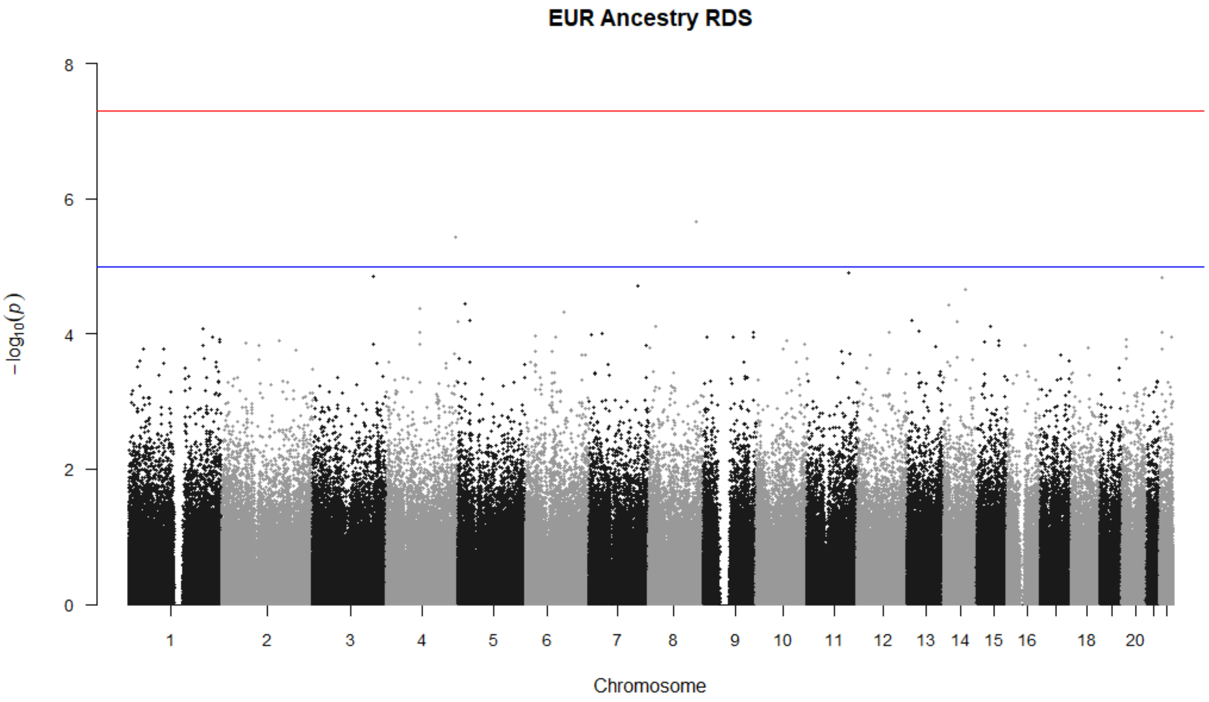
EUR Ancestry RDS
8
6
4
2
0
−log10(p)
18
20
Chromosome

d)

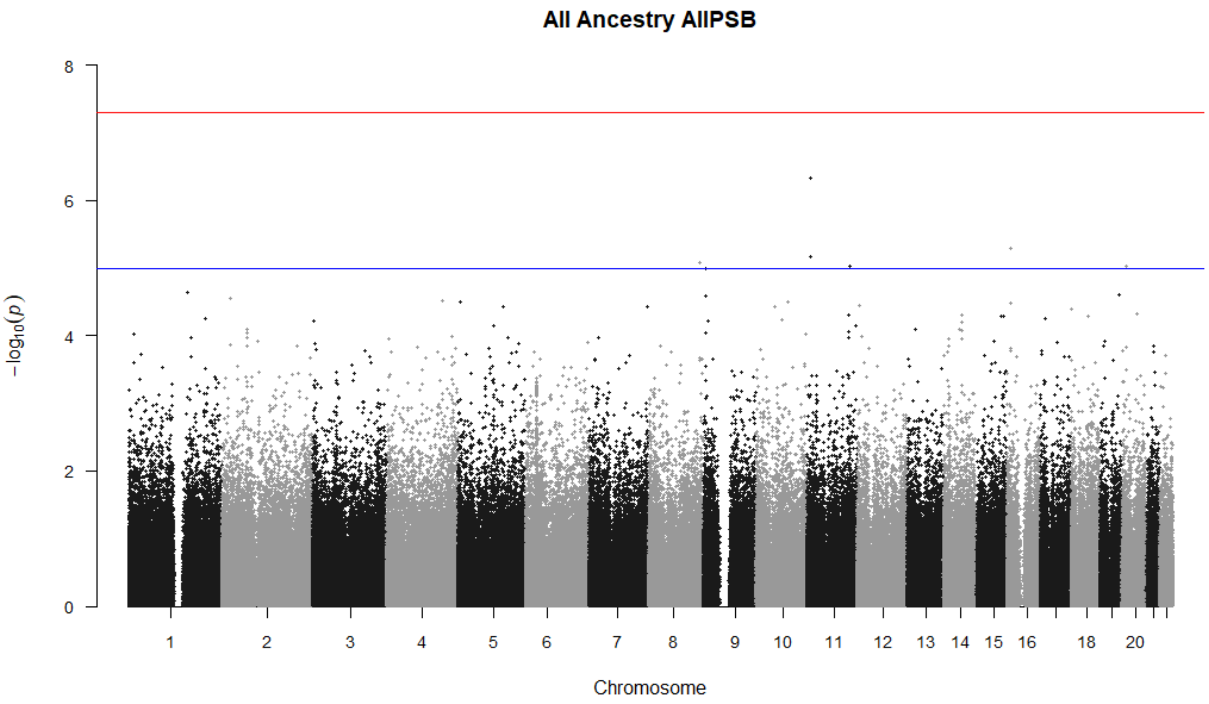
All Ancestry AllPSB
8
6
4
2
0
−log10(p)
18
20
Chromosome

e)

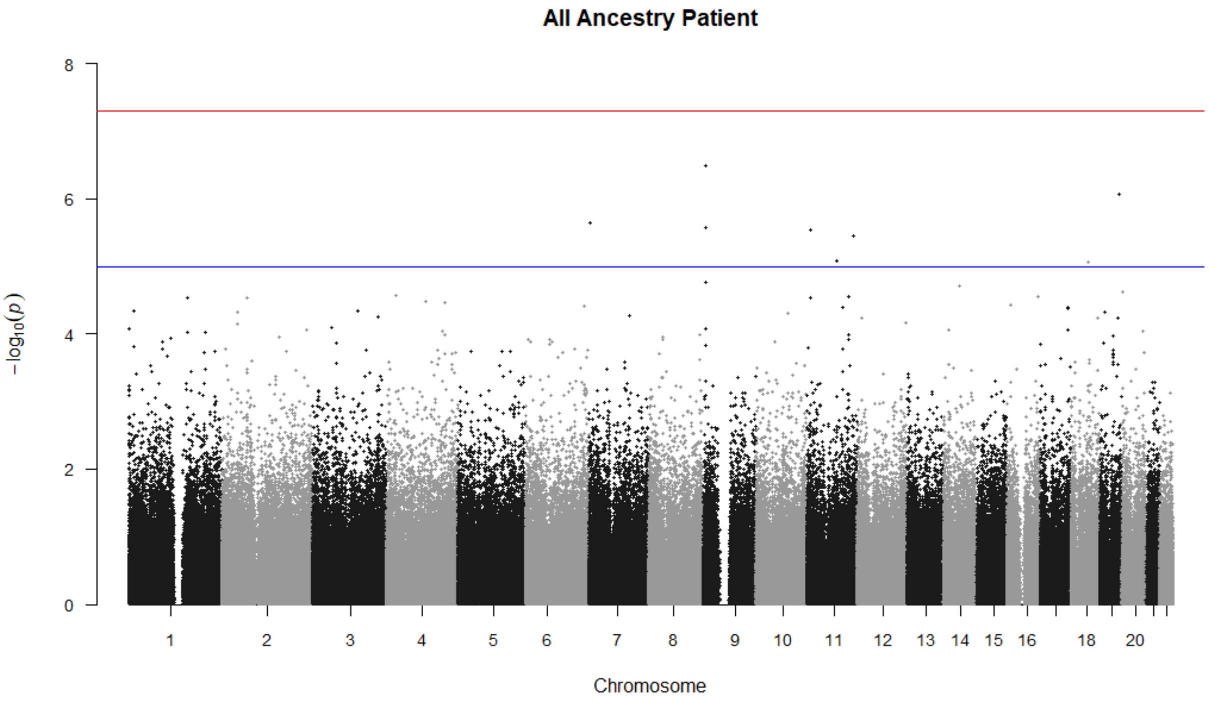

All Ancestry Patient
8
6
4
2
0
$-\log_{10}(p)$
18
20
Chromosome

f)

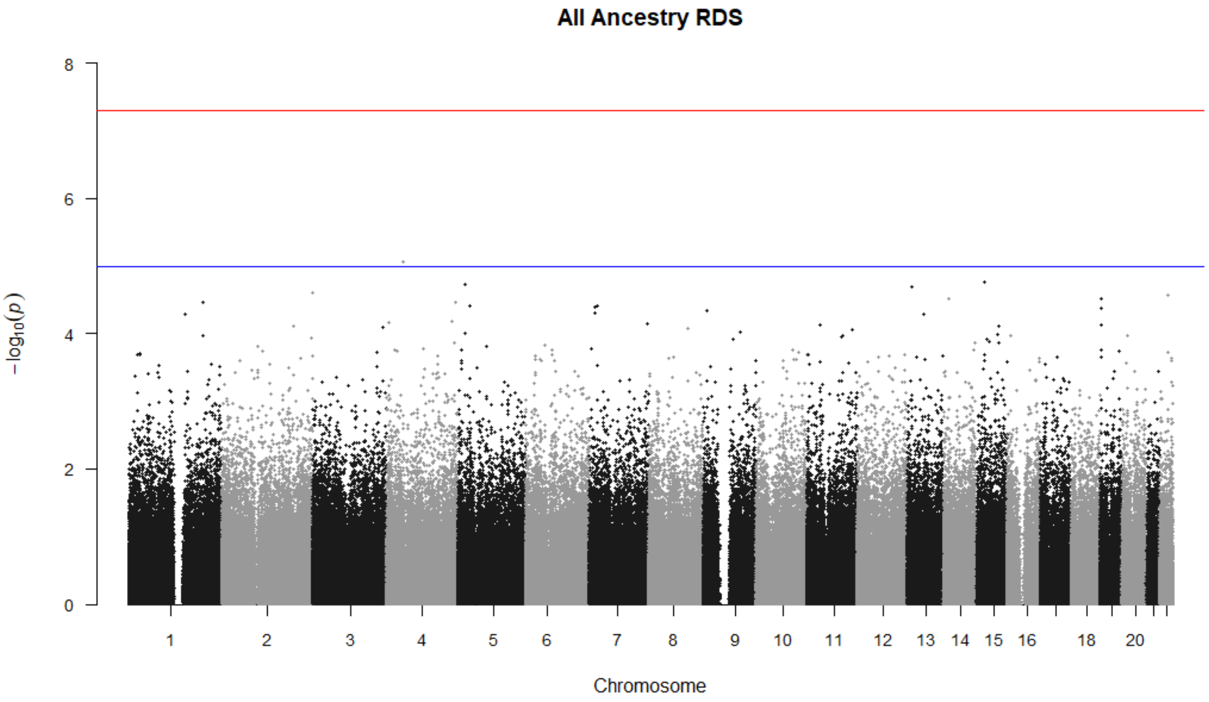

All Ancestry RDS
8
6
4
2
0
$-\log_{10}(p)$
18
20
Chromosome

**Figure S2. GTex v8 Tissue Expression Analyses** (nominal association at -log10 P Value 0.05 : 1.301). **a)** 54 Tissue Types, All-PSB, All Ancestry; **b)** General 30 Tissue Types, All-PSB, All Ancestry; **c)** 54 Tissue Types, Patient PSB, All Ancestry; **d)** General 30 Tissue Types, Patient PSB, All Ancestry; **e)** 54 Tissue Types, RDS, All Ancestry; **f)** General 30 Tissue Types, RDS, All Ancestry

**a)**

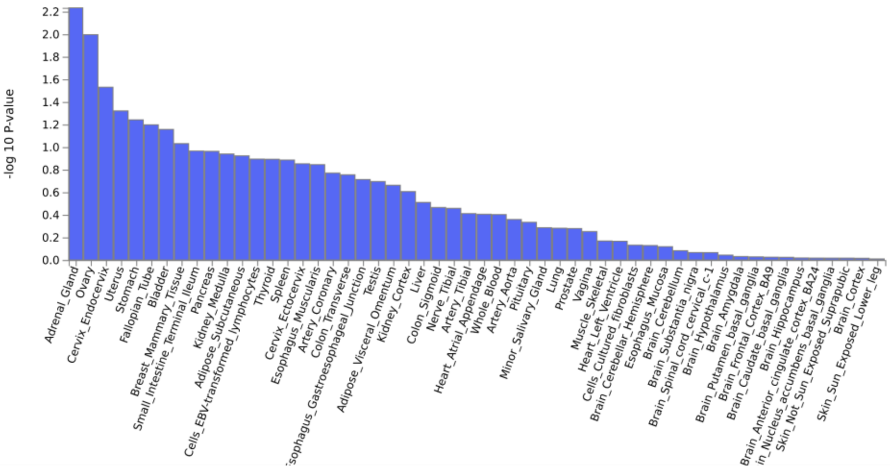

**b)**

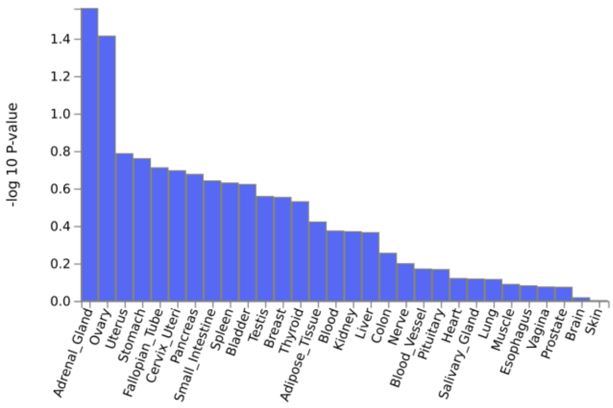

**c)**

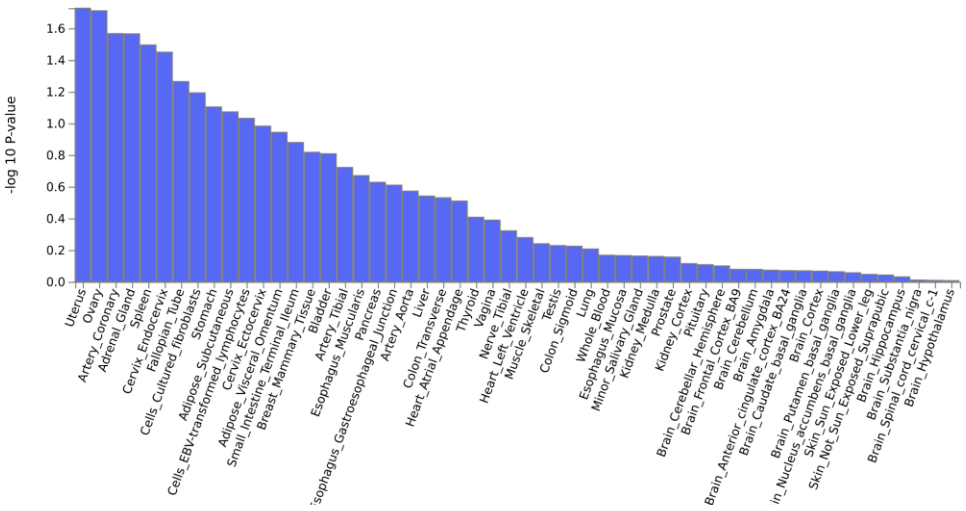

d)

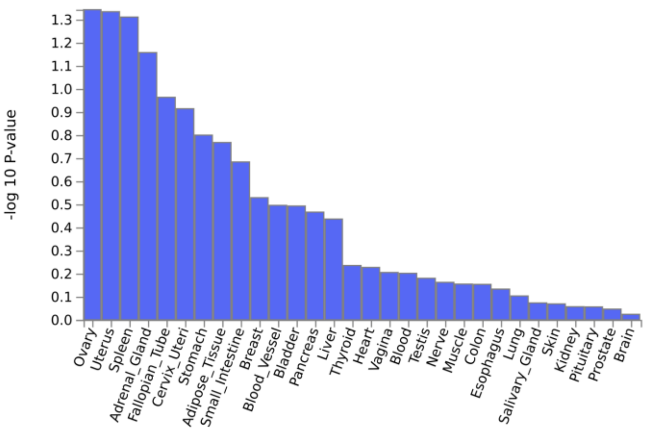
-log 10 P-value
1.3
1.2
1.1
1.0
0.9
0.8
0.7
0.6
0.5
0.4
0.3
0.2
0.1
0.0
Ovary
Uterus
Spleen
Adrenal_Gland
Fallopian_Tube
Cervix_Uteri
Stomach
Adipose_Tissue
Small_Intestine
Breast
Blood_Vessel
Bladder
Pancreas
Liver
Thyroid
Heart
Vagina
Blood
Testis
Nerve
Muscle
Colon
Esophagus
Lung
Salivary_Gland
Skin
Kidney
Pituitary
Prostate
Brain

e)

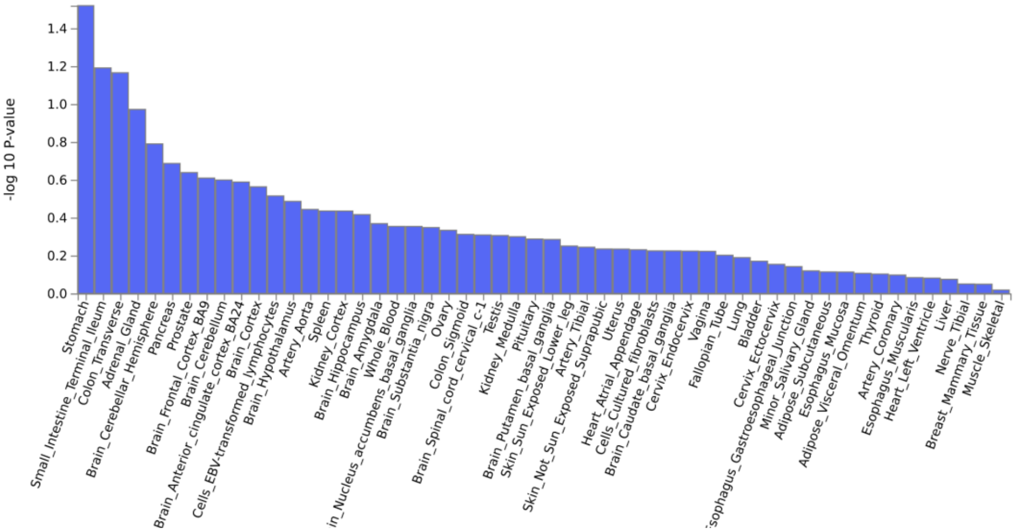
-log 10 P-value
1.4
1.2
1.0
0.8
0.6
0.4
0.2
0.0
Stomach
Small_Intestine_Terminal_Ileum
Colon_Transverse
Adrenal_Gland
Brain_Cerebellar_Hemisphere
Pancreas
Prostate
Brain_Frontal_Cortex_BA9
Brain_Cerebellum
Brain_Anterior_cingulate_cortex_BA24
Brain_Cortex
Cells_EBV-transformed_lymphocytes
Brain_Hypothalamus
Artery_Aorta
Spleen
Kidney_Cortex
Brain_Hippocampus
Brain_Amygdala
Whole_Blood
Brain_Substantia_nigra
Ovary
Colon_Sigmoid
Brain_Spinal_cord_cervical_c-1
Testis
Kidney_Medulla
Pituitary
Brain_Putamen_basal_ganglia
Skin_Sun_Exposed_Lower_leg
Artery_Tibial
Skin_Not_Sun_Exposed_Suprapubic
Uterus
Heart_Atrial_Appendage
Cells_Cultured_fibroblasts
Brain_Caudate_basal_ganglia
Cervix_Endocervix
Vagina
Fallopian_Tube
Lung
Bladder
Cervix_Ectocervix
Minor_Salivary_Gland
Adipose_Subcutaneous
Esophagus_Mucosa
Adipose_Visceral_Omentum
Thyroid
Artery_Coronary
Esophagus_Muscularis
Heart_Left_Ventricle
Liver
Nerve_Tibial
Breast_Mammary_Tissue
Muscle_Skeletal

f)

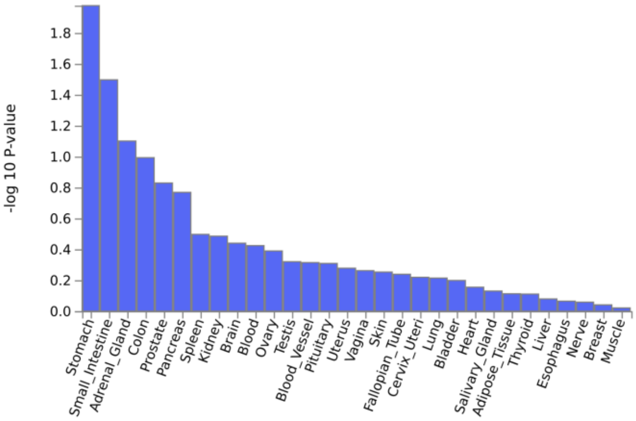
-log 10 P-value
1.8
1.6
1.4
1.2
1.0
0.8
0.6
0.4
0.2
0.0
Stomach
Small_Intestine
Adrenal_Gland
Colon
Prostate
Pancreas
Spleen
Kidney
Brain
Blood
Ovary
Testis
Blood_Vessel
Pituitary
Uterus
Vagina
Skin
Fallopian_Tube
Cervix_Uteri
Lung
Bladder
Heart
Salivary_Gland
Adipose_Tissue
Thyroid
Liver
Esophagus
Nerve
Breast
Muscle

**Figure S3. Genetic correlations between psychiatric disorders, behaviours and personality traits.** Blue denotes positive genetic correlation, while red colours denote negative correlations. Cells are blank if correlations do not meet significance threshold corrected for the number of pairwise comparisons (p=0.05/171=0.00029). AFS: Age at First Sexual Intercourse, ANX: Anxiety, ADHD: Attention Deficit Hyperactivity Disorder, BPD: Borderline Personality Disorder, ANTI: Broad Antisocial Behaviour, Extra: Extraversion, EXT: Externalising, MDD: Major Depressive Disorder OCD: Obsessive Compulsive Disorder,, OCS: Obsessive Compulsive Symptoms, NSEX: Number of Lifetime Sexual Partners, PTSD:Posttraumatic Stress Disorder, SUD: Substance Use Disorders, Open: Openness, Consc: Conscientiousness, Extra: Extraversion, Agree: Agreeableness, Neuro:Neuroticism.

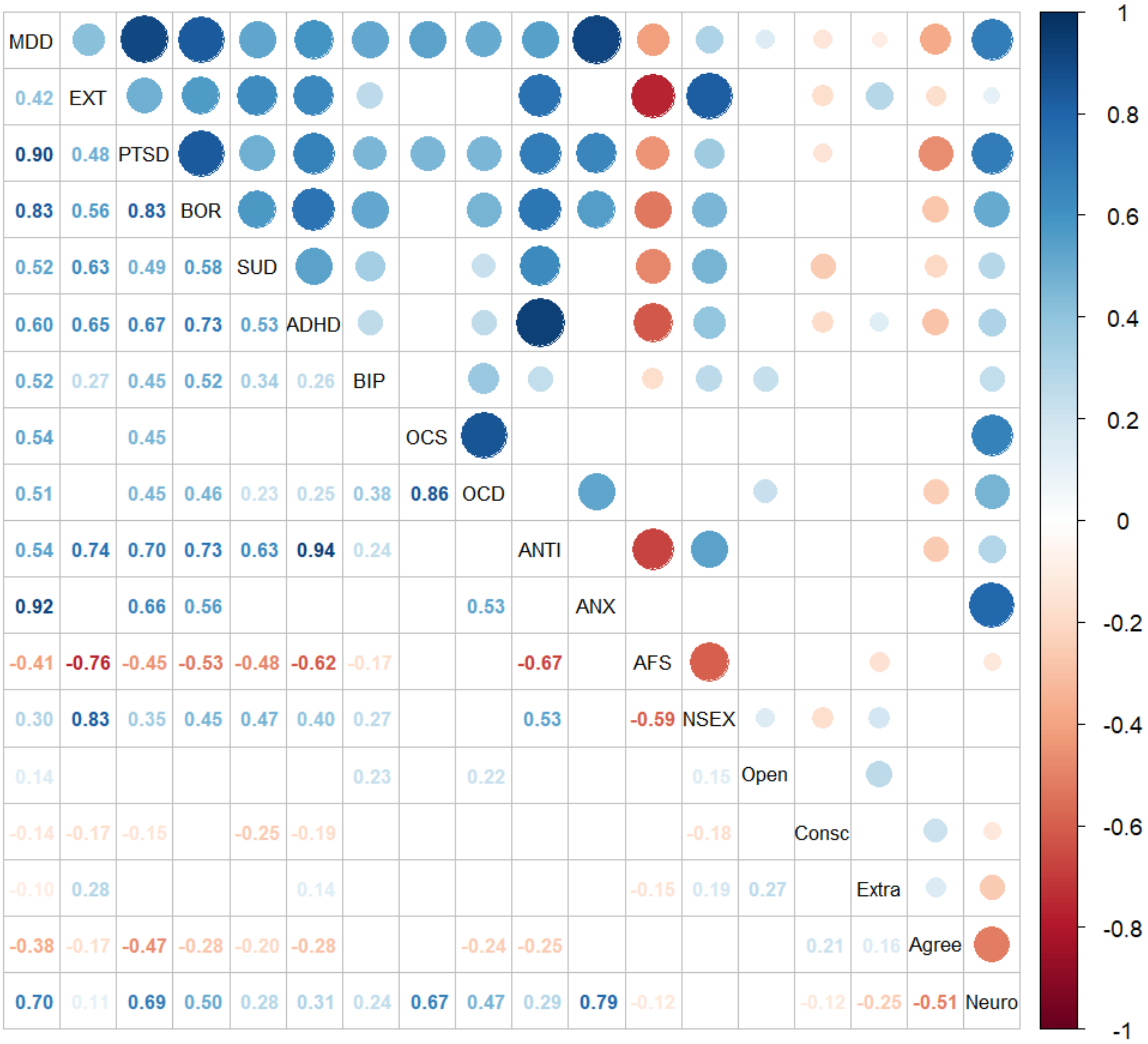

### The Behavioral Addictions Studies and Insights Consortium (BASIC)

**Kate Balestrieri, PsyD**
**Chris Chandler, MA, LMHC**
**Lauren Dummit, LMFT**
**Marcus Earle, PhD, LMFT**
**Greg Futral, PhD**
**Michelle Gaugh, MA**
**Piper Grant, PsyD, MPH**
**Alex Katehakis, PhD, MA, MFT**
**Barbara Levinson, PhD, RN, LMFT, LSOTP, CSAT Supervisor, CST Diplomate**
**Andrew Meadows, BS**
**Dan Morris, LCSW**
**Isabel Nino-de-Guzman, PhD**
**Helena Vissing, PsyD**

**Randolph Arnau, PhD***
**Bradley Green, PhD***
**Rick Isenberg, MD***
**Patrick J. Carnes, PhD***
**Katherine J. Aitchison, BM BCh, PhD, FRCPsych***

***lead investigator***

**Affiliations**

Triune Therapy Group, Los Angeles, CA, USA (KB, LD, HV)
Christian Health Group, La Jolla, CA, USA (CC)
Psychological Counseling Services, Scottsdale, AZ, USA (ME, RI)
Pine Grove Behavioral Health & Addiction Services, Hattiesburg, MS, USA (GF)
Center for Healthy Sex, Los Angeles, CA, USA (PG, AK)
Kavod Center, Rochester, NY, USA (MG, AM, DM)
Center for Healthy Sexuality, Houston, TX, USA (BL)
Gentle Path at the Meadows, Wickenburg, AZ, USA (ING, PJC)
Department of Psychology and Counseling, University of Texas at Tyler, Tyler, TX, USA (BG)
School of Psychology, University of Southern Mississippi, Hattiesburg, MS, USA (RA)
Department of Psychiatry, College of Health Sciences, University of Alberta, Edmonton, AB, Canada (KJA)
Department of Medical Genetics, College of Health Sciences, University of Alberta, Edmonton, AB, Canada (KJA)
Neuroscience and Mental Health Institute, University of Alberta, Edmonton, AB, Canada (KJA)
Women and Children's Health Research Institute, University of Alberta, Edmonton, AB, Canada (KJA)
Psychiatry Section, Division of Clinical Sciences, Northern Ontario School of Medicine, Thunder Bay, ON, Canada (KJA)
